# Lung Perfusion Disturbances Detected with MRI in Non-Hospitalized Post-COVID-19 Individuals with Dyspnea 3 -13 Months after the Acute Disease

**DOI:** 10.1101/2022.01.25.22269717

**Authors:** Jimmy Yu, Tobias Granberg, Roya Shams, Sven Petersson, Magnus Sköld, Sven Nyrén, Johan Lundberg

**Affiliations:** Department of Radiology Solna, Karolinska University Hospital, Stockholm, Sweden; Department of Molecular Medicine and Surgery, Karolinska Institutet, Stockholm, Sweden; Department of Neuroradiology, Karolinska University Hospital, Stockholm, Sweden; Department of Clinical Neuroscience, Karolinska Institutet, Stockholm, Sweden; Department of Medical Radiation Physics and Nuclear Medicine, Karolinska University Hospital, Stockholm, Sweden; Department of Respiratory Medicine and Allergy, Karolinska University Hospital, Stockholm, Sweden; Department of Medicine Solna, Karolinska Institutet, Stockholm, Sweden

**Keywords:** MeSH terms, COVID-19, Biomarkers, Dyspnea, Magnetic Resonance Imaging, Perfusion

## Abstract

**Background:** Dyspnea is a prevalent symptom in the post-COVID-19 condition, though its mechanisms are largely unknown. Lung perfusion abnormalities have been reported in acute COVID-19 and could be suspected in patients with lingering dyspnea after the acute phase.

**Objectives:** To detect pulmonary perfusion disturbances in non-hospitalized post-COVID condition with persistent dyspnea 4-13 months after the disease onset.

**Methods:** Non-hospitalized individuals reporting persistent dyspnea after COVID-19 and matched healthy controls were prospectively recruited between October 2020 and May 2021 to undergo pulmonary dynamic contrast-enhanced magnetic resonance imaging (DCE-MRI), six-minute walk test, and self-reported scales questionnaires on dyspnea and physical activity. The DCE-MRI perfusion images were quantified into two parametric values: mean time-to-peak (TTP) and TTP ratio.

**Results:** Twenty-eight persons with post-COVID condition and persistent dyspnea (mean age 46.5±8.0 years, 75% women) and 22 healthy controls (mean age 44.1±10.8 years, 73% women) were included. The post-COVID group had higher mean pulmonary TTP (0.43±0.04 vs. 0.41±0.03, P=0.011) and higher TTP ratio (0.096±0.052 vs. 0.068±0.027, P=0.032). Notably, post-COVID males had the highest values (mean TTP 0.47±0.02, TTP ratio 0.160±0.039, P<0.001 for both values compared to male controls and post-COVID females). Correlation between dyspnea and perfusion parameters was demonstrated in the males (r=0.83, P<0.001 for mean TTP; r=0.76, P=0.003 for TTP ratio), but not in females.

**Conclusions:** Lung perfusion disturbances were detected in males reporting post-COVID dyspnea using perfusion parameters from DCE-MRI. The distinct sex difference has implications for understanding the perplexing post-COVID pathophysiology and warrants future studies. DCE-MRI could provide biomarkers for such studies.

## Introduction

Post-COVID-19 condition, also known as “long-COVID”, or post-acute COVID-19 syndrome (PACS), is prevalent and can persist for months after the acute illness [1]. One prominent symptom is dyspnea, found in 24.5 % of hospitalized patients and 39.9 % in non-hospitalized patients 60 days after onset of infection [2]. In hospitalized patients, potential explanations include fibrotic-like changes visualized by computed tomography (CT), reduced spirometry volumes, and diffusion capacity [3–6]. However, a sizeable group of non-hospitalized patients also have a high incidence of dyspnea, with scarce objective findings on CT and lung function tests, but the non-hospitalized patient group is not well studied [7]. In these patients, the pathophysiological mechanisms are less clear.

During the acute SARS-CoV-2 infection, the lungs are the primary site of infection, developing into, in the worst case, an ARDS pattern [8–10]. Pulmonary involvement leads to local tissue disruption, including microvascular damage. Clinical reports support similar findings regarding an association between pulmonary hypertension and more severe disease [11]. Radiological methods including contrast-enhanced CT, dual-energy CT (DECT), and single-photon emission computed tomography combined with CT (SPECT-CT) indicate disturbances in pulmonary blood distribution [12–17]. The combined impression of unequal blood distribution in single time point scans (DECT) or composite tracer distribution (SPECT) has further support from findings in a case report of an ICU patient using dynamic contrast-enhanced magnetic resonance imaging (DCE-MRI) [18]. It has been speculated that lung perfusion disturbances could partly explain the clinical deteriorations in the acute phase [19]. The importance of examining lung perfusion in post-COVID patients has been stressed in one exploratory SPECT-CT study [20]. Indeed, early SPECT-CT studies in non-hospitalized post-COVID patients have suggested residual perfusion disturbances [21].

Lung perfusion can be studied using pulmonary angiography, DECT, SPECT/CT, and DCE-MRI. The essential advantage that sets DCE-MRI apart from the former methods is the ability to render both spatial and temporal information with a reasonable resolution, enabling detection of subtle perfusion impairments and possibly shunts. DCE-MRI is a clinically applied method for brain perfusion imaging and has also been successfully applied in lung diseases such as chronic obstructive pulmonary disease, cystic fibrosis, pulmonary embolism, pulmonary artery stenosis, and pulmonary vasculitis [22,23]. In addition, the ability to process DCE-MRI parametric maps into a few summarizing numeric values facilitates comparisons between different individuals and time points. The lack of ionizing radiation is advantageous too, especially for repeated examinations in young persons.

We hypothesized that lung perfusion disturbances might exist within this patient group and contribute to dyspnea. Therefore, in this prospective study, we applied DCE-MRI to investigate if pulmonary perfusion disturbance exists in non-hospitalized post-COVID individuals and whether it might be associated with dyspnea.

## Methods

### Ethical considerations

The Regional Ethics Review Board in Stockholm and the Swedish Ethical Review Authority approved this prospective cross-sectional study performed October 2020 and May 2021 (original approval number 2018/2416-31 with amendments 2020-00047, 2020-02535, 2021-00815). Written informed consent was obtained from all participants.

### Participant selection and enrollment

Inclusion criteria for the post-COVID condition group was a history of past COVID-19 infection, verified by real-time polymerase chain reaction (rt-PCR) and persistent dyspnea at enrollment. Participants were recruited through a patient network in Sweden and kindly asked to recruit an age and sex-matched healthy control, if possible. Controls were included if they had a negative antibody test within three weeks from the imaging point and the absence of COVID-suspect symptoms since the pandemic started.

Exclusion criteria for both groups were (i) a history of smoking for more than five years, (ii) any cardiovascular or (iii) pulmonary conditions requiring medical follow-up or treatment. In addition, all participants were asked to fill out an MRI safety checklist, and any contraindications meant exclusion from the study.

One male person with idiopathic pulmonary fibrosis [24], diagnosed by the Respiratory medicine clinic at Karolinska University Hospital, was additionally included as a positive control. He was in his 80s (more than 4 SD older than the post-COVID group) but with a BMI of 23,8 (within 1 SD). His disease severity was assessed as “mild to moderate” [25] based on a forced vital capacity of 92 %, predicted and diffusion capacity of 63 %, and did not require long-term oxygen treatment. Computed tomography showed subpleural reticular changes with traction bronchiectasis and no honeycombing. The total volume of morphologic changes was visually assessed to be 10-20% of the total lung volume.

### Clinical data acquisition

All clinical data were collected directly in conjunction with the MRI imaging session. Essential patient characteristics were recorded: age, sex, body height, and body weight from which the body mass index (BMI) was calculated, date of disease onset, and confirmed rt-PCR test result.

Dyspnea severity was quantified through two validated self-reported symptom scales: the modified Medical Research Council dyspnea scale (mMRC) and Chronic Obstructive Pulmonary Disease assessment test (CAT) [26,27]. The CAT scale includes several questions regarding different symptoms related to COPD. Only one question, the one related to exertional dyspnea, was used in the analysis. In both scales, a higher number means more dyspnea.

Subjective exertional impairment was quantified through the validated self-reported Frändin-Grimby [28]. A lower number means less daily physical activity.

Objective exertional impairment was quantified through a 6-min walking test (6MWT), performed according to the American Thoracic Society 2002 guidelines by J.Y. (pulmonologist and fifth-year radiology resident) [29]. We also related the absolute walking distance to expected values based on normative data [30] to account for differences in age, body height, and body weight which might affect the absolute walking distance.

### MRI image acquisition

Pulmonary imaging was performed by J.Y. and R.S. (MRI technologist) under the supervision of T.G. (radiologist) on a Siemens MAGNETOM Skyra 3 Tesla MRI scanner (Siemens Healthineers, Erlangen, Germany). The imaging protocol was designed by J.Y., T.G., S.P. (MRI physicist), and J.L. (radiologist), including three morphological sequences and one perfusion sequence. Prior to entering the scanner, the patient was given breath-hold training. The entire imaging session lasted 15-20 minutes.

Three morphological image non-enhanced sequences, each performed during inspiration breath-hold for 16 seconds, were used to identify lung opacifications: a coronal 2D T2-weighted half-Fourier single-shot spin-echo (“HASTE”, field-of-view 400×400 mm, voxel size 2.1 × 2.1 × 5 mm, echo/repetition times at 23/400 ms, 36 slices); a transverse 2D T1-weighted spoiled gradient echo (“VIBE”, field-of-view 262×400 mm, voxel size 1.0 × 1.0 × 4 mm, flip angle 5 °, echo/repetition times at 1.9/4 ms, 72 slices); a coronal 3D ultrashort echo-time spiral VIBE (field-of-view 600×600 mm, voxel size 2.3 × 2.3 × 2.3 mm, flip angle 5 °, echo/repetition times at 0.05/2.62 ms, 104 slices).

The DCE-MRI was acquired through a 4D time-resolved MRI angiography sequence with a keyhole T1-weighted gradient-recalled-echo (“TWIST”, field-of-view 450 × 450 mm, voxel size 1.5 × 1.5 × 4 mm, echo/repetition times at 0.64/1.9 ms, 26 slices, a total of 90 phases in 40 seconds). A Spectris Solaris EP contrast injector (MEDRAD, Pittsburgh, USA; Bayer, Leverkusen, Germany) was used to administer gadoterate contrast agent (Clariscan, GE Healthcare, Chicago, USA), 0.5 mmol/ml, 2 ml, followed by 20 ml 0.9% saline solution with five ml/s. The patients were told to inhale and hold their breath for as long as possible during the 40-second image acquisition, preceded by ten deep breaths.

Repeatability was evaluated by performing DCE-MRI twice on two consecutive days in one healthy control.

### MRI image assessment and quantification

Morphological imaging was evaluated by J.Y. and independently verified by J.L. or S.N. (radiologists). MRI perfusion series were analyzed using MATLAB (version R2020b, The Mathworks Inc., Natick, USA) as previously described using an in-house developed post-processing pipeline [18]. Briefly, lungs were manually masked, and regions of interest (ROI) were manually selected in the pulmonary artery (PA) before its bifurcation and in the aortic arch at its superior portion. This was performed by J.Y. and independently verified by J.L., blinded to clinical data. To allow for a structured and quantitative comparison between individuals and groups, the perfusion scans were normalized in the time domain to the peaks of the PA (as 0) and aorta (as 1) and summarized using two numeric parameters: Time-to-peak (TTP) ratio and mean TTP. TTP parametric maps were calculated using the MATLAB function max. Temporal smoothing with a robust Locally Weighted Scatterplot Smoothing algorithm was applied before TTP calculation. Mean TTP was calculated as the mean of lung TTP values between PA and aorta. TTP ratio was calculated as the fraction of voxels with TTP values later than the aorta, i.e., a bolus arrival after the aorta.

### Statistics analysis

All values reported are the mean and standard deviations unless explicitly stated otherwise. Categorical variables were evaluated with the Pearson chi-square test, and continuous variables were assessed with the Students t-test, while ordinal variables were evaluated with the Mann-Whitney test. When analyzing sex differences, ANOVA with Dunn-Sidak corrections for multiple comparisons was used since there is an uneven number of individuals to compare between multiple groups. Finally, correlations were assessed with the Spearman correlation method. Statistical analyses were performed using MATLAB (version R2020b, The Mathworks Inc., Natick, USA). To generate Figures 3 and 4, curves were smoothed using the moving mean method with a smoothing factor of 0.2, then interpolated using the modified Akima method. This was done to generate a unified x-axis for simultaneous plotting. No interpolation was performed prior to calculating the mean TTP and TTP ratio. Normalized TTP-distribution curves from the two MRI scans performed in one healthy control were compared, with the difference between these two curves expressed as the mean coefficient of variation.

## Results

In total, 51 participants were recruited. Included in the data analysis were 28 individuals with longstanding symptoms and RT-qPCR positive testing at the time of disease, 22 healthy controls, and 1 positive control with idiopathic pulmonary fibrosis. For brevity, the RT-qPCR positive group with longstanding symptoms following COVID-19 will be referred to as the post-COVID group. In addition, 1 patient with idiopathic pulmonary fibrosis was included as a positive control. None in the post-COVID group had a history of being hospitalized or receiving treatment during the acute phase. All participants tolerated the image session well. A flow chart of the study inclusions is presented in Figure 1. The two cohorts had no significant difference in neither mean age, the proportion of female participants, mean weight, nor mean BMI, as detailed in Table 1. In the post-COVID group, the mean time from symptom onset to the exam was 7.7±3.6 months, with a biphasic distribution. About one-half of the post-COVID group performed imaging 4-6 months after symptom debut, and the rest 10-13 months after, reflecting the first and second wave of COVID-19 pandemic in Sweden.

**Table 1:**
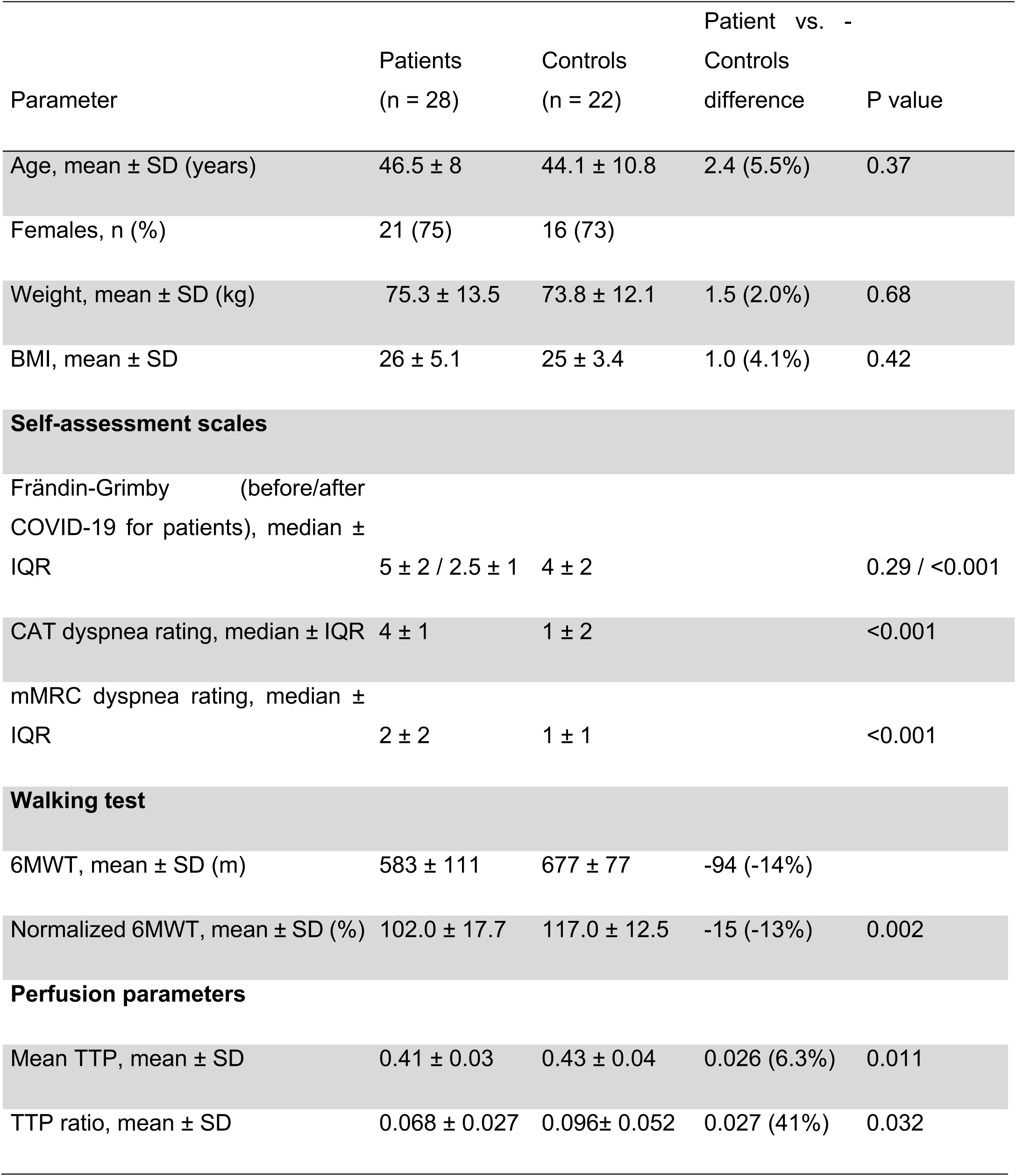
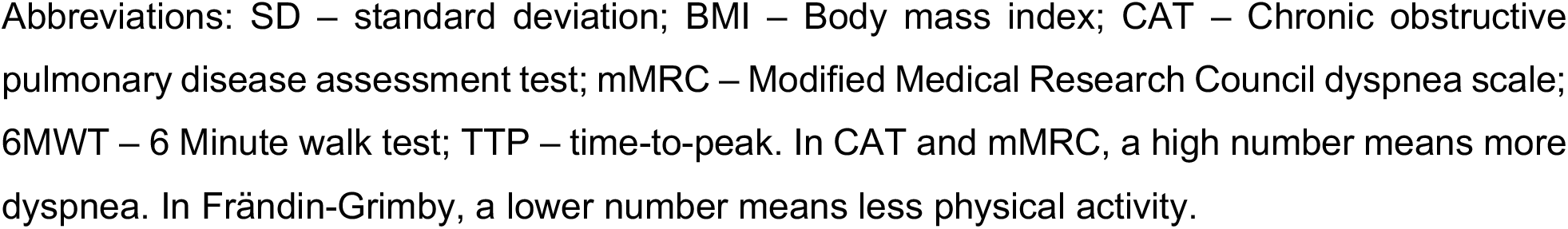
Patient characteristics, clinical parameters and perfusion results.

| Parameter | Patients<br>(n = 28) | Controls<br>(n = 22) | Patient vs. Controls<br>difference | P value |
| --- | --- | --- | --- | --- |
| Age, mean ± SD (years) | 46.5 ± 8 | 44.1 ± 10.8 | 2.4 (5.5%) | 0.37 |
| Females, n (%) | 21 (75) | 16 (73) |  |  |
| Weight, mean ± SD (kg) | 75.3 ± 13.5 | 73.8 ± 12.1 | 1.5 (2.0%) | 0.68 |
| BMI, mean ± SD | 26 ± 5.1 | 25 ± 3.4 | 1.0 (4.1%) | 0.42 |
| **Self-assessment scales** |  |  |  |  |
| Frändin-Grimby (before/after COVID-19 for patients), median ± IQR | 5 ± 2 / 2.5 ± 1 | 4 ± 2 | | 0.29 / <0.001 |
| CAT dyspnea rating, median ± IQR | 4 ± 1 | 1 ± 2 | | <0.001 |
| mMRC dyspnea rating, median ± IQR | 2 ± 2 | 1 ± 1 | | <0.001 |
| **Walking test** |  |  |  |  |
| 6MWT, mean ± SD (m) | 583 ± 111 | 677 ± 77 | -94 (-14%) | |
| Normalized 6MWT, mean ± SD (%) | 102.0 ± 17.7 | 117.0 ± 12.5 | -15 (-13%) | 0.002 |
| **Perfusion parameters** |  |  |  |  |
| Mean TTP, mean ± SD | 0.41 ± 0.03 | 0.43 ± 0.04 | 0.026 (6.3%) | 0.011 |
| TTP ratio, mean ± SD | 0.068 ± 0.027 | 0.096 ± 0.052 | 0.027 (41%) | 0.032 |

Abbreviations: SD – standard deviation; BMI – Body mass index; CAT – Chronic obstructive pulmonary disease assessment test; mMRC – Modified Medical Research Council dyspnea scale; 6MWT – 6 Minute walk test; TTP – time-to-peak. In CAT and mMRC, a high number means more dyspnea. In Frändin-Grimby, a lower number means less physical activity.

**Figure 1.**
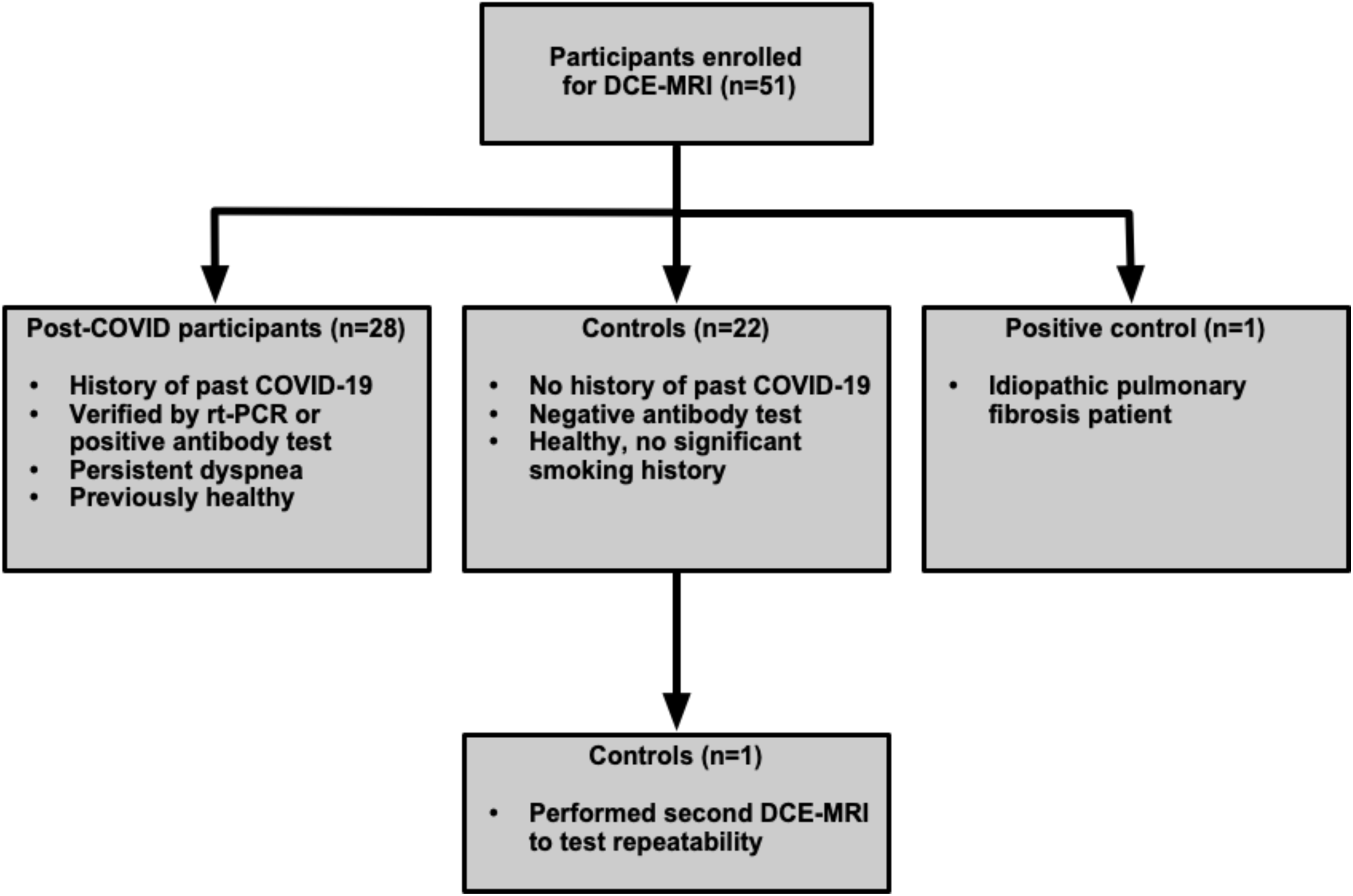
Abbreviations: DCE-MRI – dynamic contrast-enhanced magnetic resonance imaging

The post-COVID group had a lower current self-reported physical activity (2.5±1) than the control group (4±2, P<0.001) according to Frändin-Grimby, but the re-called physical activity (5 ± 1) before COVID-19 was not significantly different (P=0.29) from the controls. The post-COVID group also reported more dyspnea on CAT (4±1 vs. 1±2, P<0.001) and mMRC (2±2 vs. 1±1, P<0.001).

When performing 6MWT, the post-COVID group had a shorter walking distance, both in actual meters (583±111 vs. 677±77, P=0.001) and normalized value, as a percentage of the expected reference value (102.0±17.7 % vs. 117.0±12.5 %, P=0.002). However, both groups remained within the clinically normal range.

No systematic structural changes were identified on the morphological lung MRI imaging. Only small atelectatic opacities (max. 7 mm) in 4/28 (14%) participants in the post-COVID group were visualized. None of the controls had any detectable lung changes. There was no correlation between time from symptom onset to MRI and any of the perfusion parameters.

On the TTP-maps, there was visually a late bolus arrival in many post-COVID participants, exemplified in Figure 2. The TTP ratio, capturing bolus arrival later than the aortic peak in the lung parenchyma, was significantly higher in the post-COVID group at 0.096±0.052 vs. 0.068±0.027 in the control group (P=0.032). The mean TTP was significantly higher in the post-COVID group at 0.43±0.04 vs. 0.41±0.03 (P=0.011). A higher mean TTP indicates an overall slower inflow of contrast bolus. Notably, the post-COVID group displayed a more considerable within-group variability regarding both TTP ratio and mean TTP (Figure 3). The group differences are also reported in Table 1.

**Figure 2.**
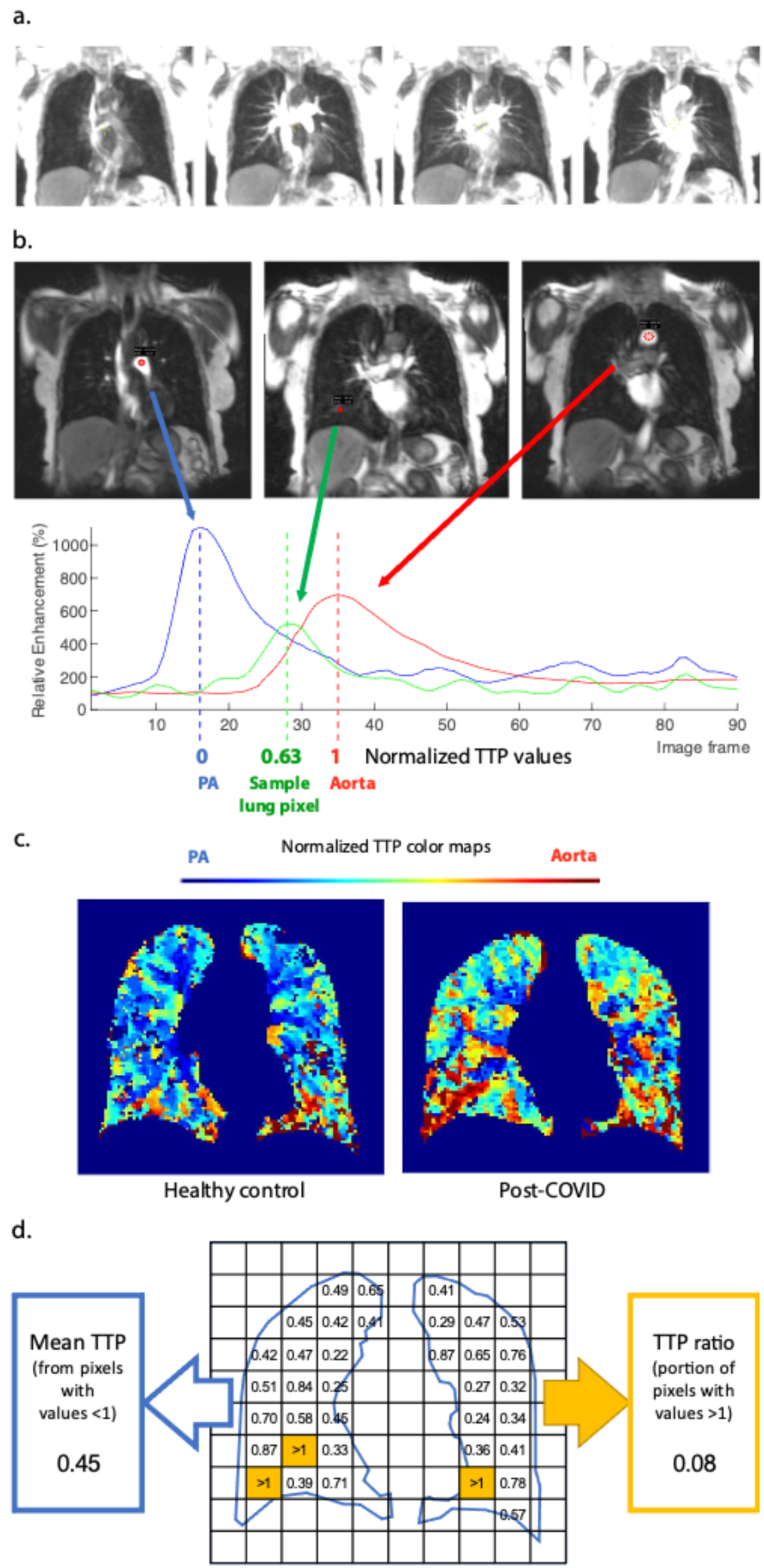
*Post-processing pipeline of the perfusion series* a. Coronal slices from the DCE-perfusion series with the same anatomical position and window setting; from left to right: before contrast arrival, pulmonary artery-peak, parenchymal phase, aortic peak. b. Pictorial explanation of normalized TTP map creation. ROI:s (red circles) were placed in the pulmonary artery and aorta. Relative enhancement was plotted against time to find the respective enhancement peak. The lung pixel TTPs time curve was then normalized to the timing of the TTP peaks in the pulmonary artery (0) and aorta (1). c. Sample coronal slice of lung masked TTP map for healthy control (left) and post-COVID participant (right). The colors represent normalized TTP values (PA/0 as blue, aorta/1 as red). d. Calculation of summarizing parameters. Note: numbers and the exaggerated pixel size are for illustrations purposes and do not mirror the actual data.

**Figure 3.**
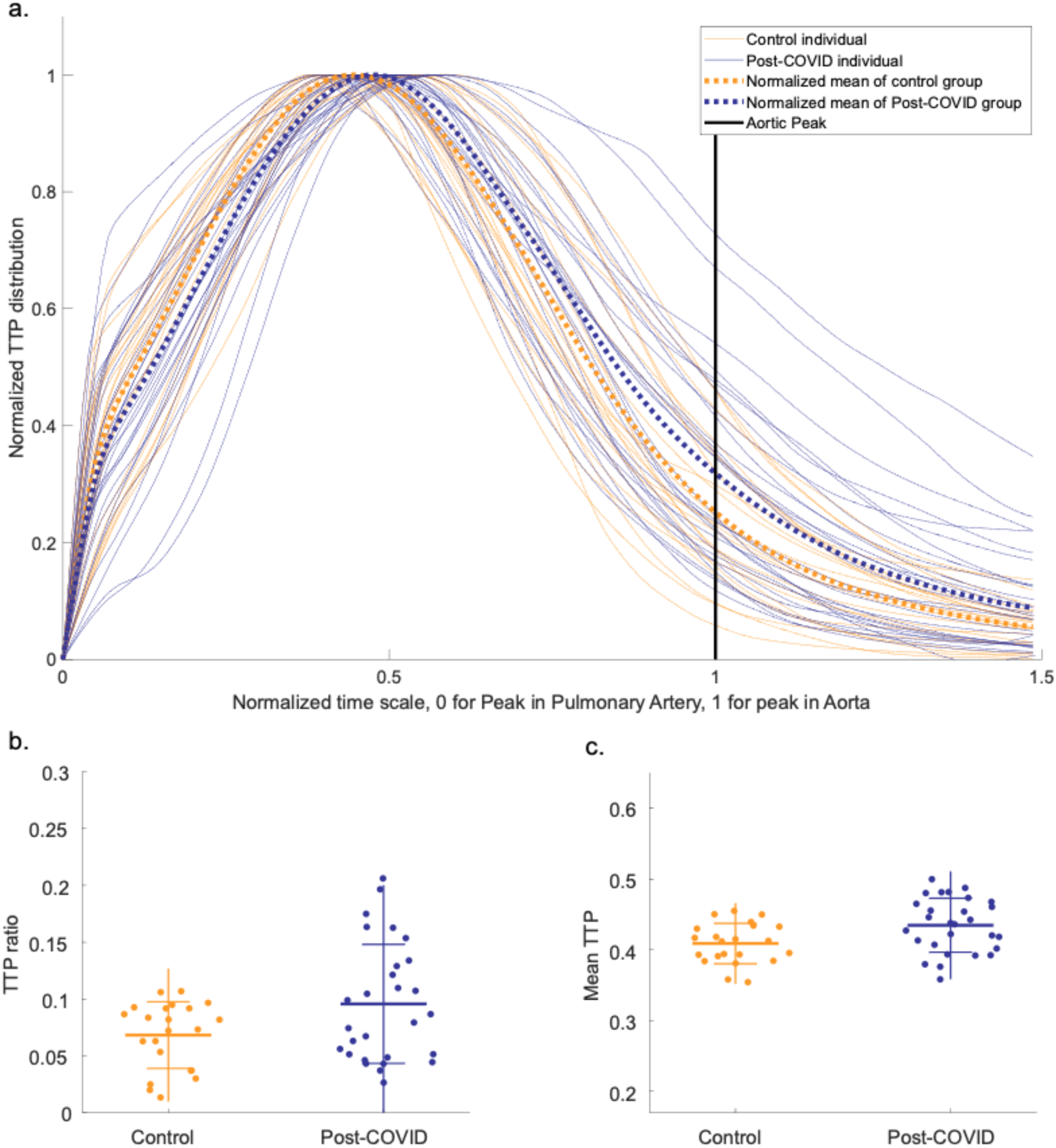
*Perfusion distribution curves* a. Individual TTP distribution curves, Y-normalized by its intensity peak. Mean curve for each group in thick dotted lines. b. TTP ratio of the respective group, representing the area-under-curve portion after the aortic peak c. Mean TTP of the respective group.

Sub-group analyses of a possible sex difference revealed that post-COVID males had worse mean TTP (0.47±0.03 vs. 0.40±0.03, P=0.001) and TTP ratio (0.160±0.039 vs. 0.082±0.027, P=0.001) compared to the male control group. The comparisons are summarized in Figure 4. The female post-COVID group did not differ significantly from the female control group, neither regarding mean TTP nor TTP ratio. Notably, the female post-COVID participants displayed greater variability in perfusion metrics relative to the female controls (Figure 4).

**Figure 4.**
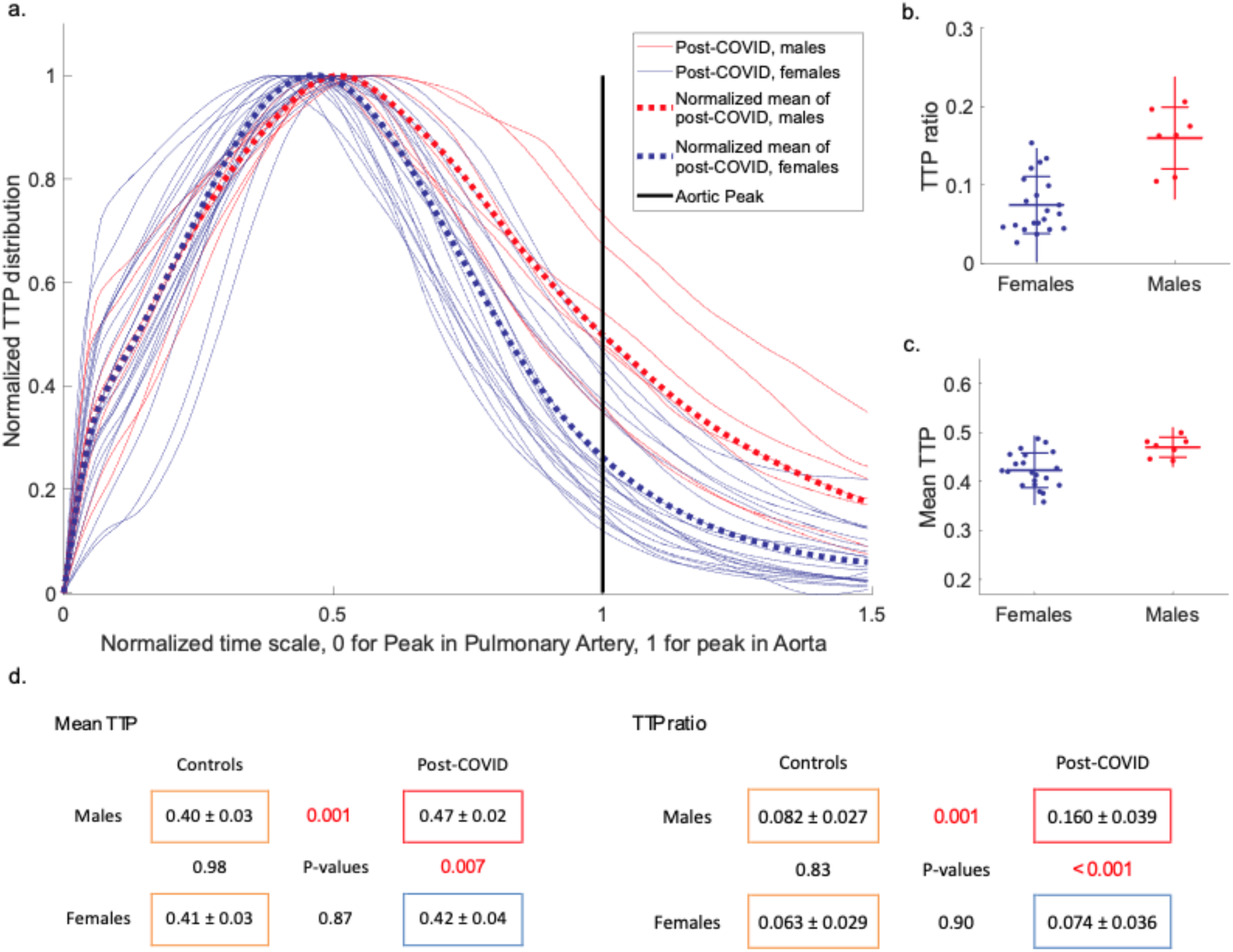
*Sex differences in perfusion metrics* a. Individual TTP distribution curves for the post-COVID males and females, respectively, Y-normalized by its intensity peak. Mean curve for each group in thick dotted lines. b. TTP ratio of the respective group, representing the area-under-curve portion after the aortic peak c. Mean TTP of the respective group.

The overall correlation between self-reported dyspnea, 6MWT and TTP values are visualized as scatterplots (Figure 5). Analyzing all participants, we found a correlation between CAT and mean TTP (r = 0.35, P=0.013). For males, this correlation was even more pronounced, both between CAT and mean TTP (r = 0.83, P=< 0.001), and between CAT and TTP ratio (r = 0.76, P=0.003). Results from 6MWT did not correlate to MRI-DCE parameters.

**Figure 5.**
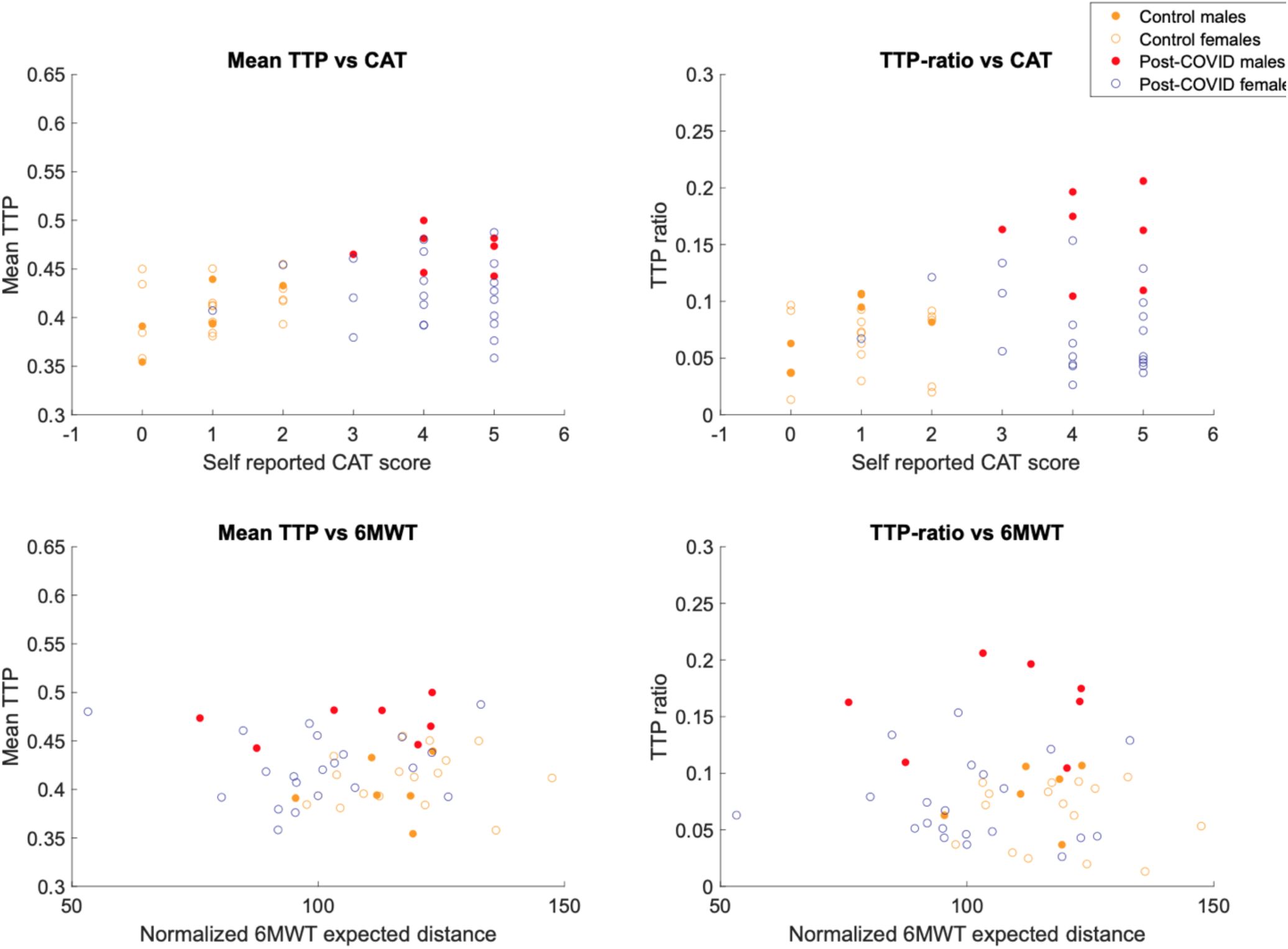
*Scatter plots for perfusion values and clinical assessments* Abbreviations: TTP – Time-To-Peak, CAT - Chronic obstructive pulmonary disease assessment test, mMRC – Modified Medical Research Council dyspnea scale. In CAT and mMRC, a high number means more dyspnea. In Frändin-Grimby, a lower number means less physical activity

To put the degree of perfusion impairment of the post-COVID group into a clinical perspective, one male patient in his 80s with idiopathic pulmonary fibrosis was included in the study as a positive control. The walking distance was 440 m (87% of the expected reference value). The TTP-mean for this patient was 0.45 and the TTP-ratio 0.15, which is comparable to the most pathological values seen in the post-COVID group. This patient also had subtle peripheral morphologic changes involving a larger lung volume compared to the worst post-COVID participants.

One healthy control performed DCE-MRI on two consecutive days to further characterize the DCE sequence and post-processing variability. The mean TTP was 0.39 and 0.38, TTP-ratio was 0.0248 and 0.0238, respectively, over the two sessions (Figure 6).

**Figure 6.**
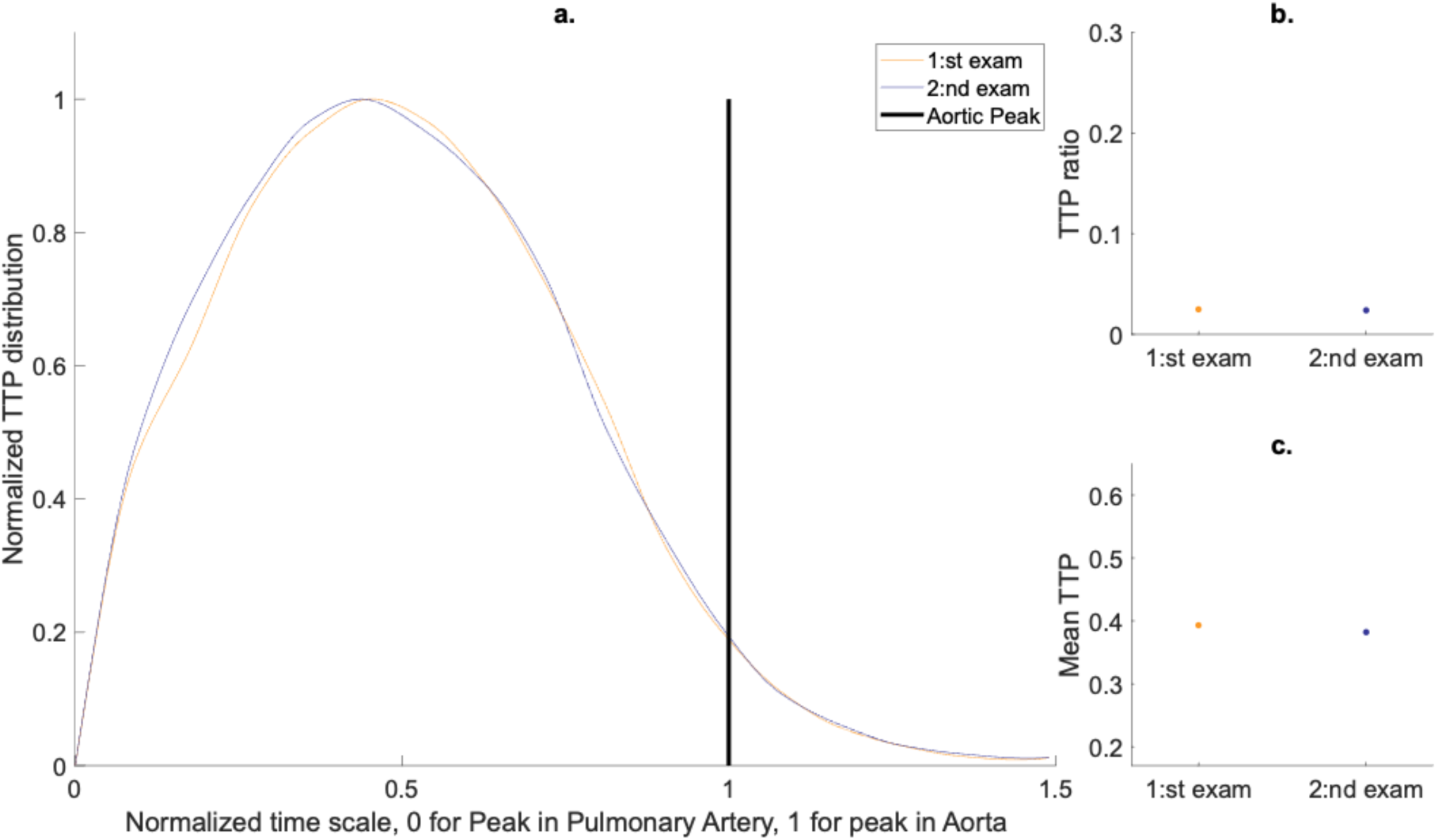
*Repeatability in one healthy control* a. TTP distribution curves from 2 consecutive days (mean coefficient of variation 3.7%) b. TTP ratios were 0.0248 and 0.0238 for the respective scans (4.0% difference) c. Mean TTPs were 0.393 and 0.382 for the respective exam (2.8% difference)

Diaphragm movement during the DCE sequence was evaluated for all participants as a possible source of variability. The reference point for the movement was the topmost point of the right diaphragm in the mid-coronal slice (slice 13 of 26). The movement was quantified in the number of voxels during the bolus transit. Six participants had a diaphragm movement exceeding two voxels (3 mm), and the most considerable movement during bolus passage was six voxels (9 mm). Most (86%) of the participants could hold their breath during the entire 40 seconds image acquisition. Seven subjects (14%) resumed breathing in the latter half, well after bolus transit.

## Discussion

The high prevalence of post-COVID dyspnea is developing into a potentially sizeable clinical and socioeconomic issue. However, in non-hospitalized patients, objective findings are scarce, and the mechanisms are largely unknown. Lung perfusion disturbances are a significant pathophysiologic finding in acute COVID-19 infection. We, therefore, wanted to investigate if perfusion disturbances persist in post-COVID. Applying DCE-MRI, we detected lung perfusion disturbances in a prospective convenience sample of non-hospitalized post-COVID patients. In males, there was a significant correlation with dyspnea.

Sex differences in pathophysiology have been reported regarding acute COVID-infection. Men have a higher expression of angiotensin-converting enzyme II-receptor, abundant in the lung, and a higher propensity for COVID-associated respiratory failure and mortality [31]. The post-COVID perfusion sex difference could thus be anticipated. Notably, in our study, post-COVID males had distinctively worse perfusion parameters than healthy male controls and female counterparts. The lack of a stronger correlation between perfusion and clinical parameters in females could partly be expected, given that perfusion abnormalities were less pronounced among females. We should point out that our results do not exclude the possibility of lung perfusion disturbances in females. We observed considerable variability in the female group, with some individuals showing impaired perfusion comparable to the males. One possible explanation is that, in our convenience sample, we have recruited a more heterogeneous female population with other underlying causes for their self-reported dyspnea.

An essential factor to discuss is that the imaging was conducted during rest and not exertion. Pulmonary perfusion is directly linked to cardiac output, which can increase many-fold during heavy physical exertion. In addition, the lung perfusion becomes more heterogeneous as pulmonary blood flow increases [32]. This increase might be more pronounced in post-COVID individuals. Due to practical reasons, it is challenging to perform DCE-MRI during physical exertion, which applies to other perfusion imaging methods as well. A possible extension of the current work could be pharmacologically induced stress-testing during DCE-MRI, thereby highlighting possible group differences.

Based on our results, we believe this lung DCE-MRI method is feasible for exploring lung perfusion in a larger clinical setting, applicable to any MRI scanner. When done in conjunction with other diagnostic and interventional trials, the underlying cause of perfusion disturbances may be further elucidated, be it microthrombi, parenchymal destruction, dysregulation, or secondary to ventilation disturbances. DCE-MRI was chosen as our study method for its many advantages, including temporal information and lack of radiation. Its application in lung perfusion imaging in other conditions has thus far yielded convincing results [23]. Analysis of TTP parametric maps in high-altitude pulmonary edema susceptible individuals have previously been reported [33]. The post-processing method whereby the TTP maps are normalized in the time domain and summarized into mean TTP and TTP ratio is straightforward and has been successfully applied in one patient treated with intensive care for COVID-19 and a porcine model simulating severe COVID-19 [18].

To our knowledge, this is the largest cohort examined using lung DCE-MRI in COVID-19 and post-COVID. These patients did, by definition, not receive any medical treatment during their acute phase, possibly representing a “mild” stratum of patients. Our findings thus indicate a possible physiological disturbance beyond morphological changes, similar to other studies using DECT or hyperpolarized ^129^Xe MRI [6,34]. What is surprising is our finding of perfusion disturbances up to one year after symptom onset. This finding could suggest a long-lasting or even perpetual lung injury in some patients. Further studies would elucidate the factors predicting long-term outcomes.

This study has some limitations, including a lack of other supporting imaging modalities. CT scans would likely detect morphological lung changes with higher sensitivity, but the current MRI protocol was optimized with advanced morphological imaging, including ultra-short echo time imaging that has recently been successfully applied in cystic fibrosis [35]. This study can be regarded as exploratory on the potential use of DCE-MRI in post-COVID, and future studies should combine multiple imaging modalities such as CT, echocardiography, and pulmonary physiological measurements such as spirometry. Nevertheless, we detect heterogeneity in our post-COVID group, indicative of a significant systematic difference on the group level. The heterogeneity supports a possible perfusion problem in the post-COVID group. A technical limitation that is hard to mitigate is the possible contribution of inspiration during breath-hold. A higher degree of inspiration can increase pulmonary resistance by stretching alveolar capillaries and by vessel compression due to higher intrathoracic pressure [36]. Nevertheless, the repeatability of a healthy volunteer was excellent. In addition, the unstructured recruitment of patients through a network of self-identified “long haulers” following COVID-19 could be considered both a weakness and a strength. The potentially increased heterogenicity is offset by a better reflection of clinical reality. In hindsight, with regards to perfusion disturbances detected predominantly in men, the male participants would have been more numerous. It could be argued that the clinical tests we used, originally meant for debilitating lung diseases, were not an optimal match for our participants. In addition, *post hoc* assessments of physical activity before COVID-19 may also be subject to a recall bias. Nevertheless, our results suggest that perfusion impairment contributes to dyspnea and reduced exercise capacity in our sample of non-hospitalized post-COVID individuals.

In conclusion, by calculating the mean TTP and TTP ratio from DCE-MRI, perfusion disturbances can be detected in non-hospitalized patients long after an acute COVID-19 infection. This method could investigate other patient groups, the most salient being hospitalized, post-COVID patients. Our results regarding sex differences are important to consider for future studies addressing the role of lung perfusion disturbances in post-COVID pathophysiology.

## Data Availability

The anonymized datasets generated and analyzed in the study are available from the corresponding author on reasonable request.

## Acknowledgments

The authors are grateful to the study participants for providing their time.

## References

1. Soriano JB, Murthy S, Marshall JC, Relan P, Diaz JV, Condition WCCDWG on P-C-19. A clinical case definition of post-COVID-19 condition by a Delphi consensus. Lancet Infect Dis 2021.

2. Fernández-de-las-Peñas C, Palacios-Ceña D, Gómez-Mayordomo V, et al. Prevalence of post-COVID-19 symptoms in hospitalized and non-hospitalized COVID-19 survivors: A systematic review and meta-analysis. Eur J Intern Med 2021; 92: 55–70.

3. Huang C, Huang L, Wang Y, et al. 6-month consequences of COVID-19 in patients discharged from hospital: a cohort study. Lancet 2021; 397: 220–32.

4. Han X, Fan Y, Alwalid O, et al. Six-Month Follow-up Chest CT findings after Severe COVID-19 Pneumonia. Radiology 2021; 299: 203153.

5. Li H, Zhao X, Wang Y, et al. Damaged lung gas exchange function of discharged COVID-19 patients detected by hyperpolarized 129Xe MRI. Sci Adv 2021; 7: eabc8180.

6. Grist JT, Chen M, Collier GJ, et al. Hyperpolarized 129Xe MRI Abnormalities in Dyspneic Participants 3 Months after COVID-19 Pneumonia: Preliminary Results. Radiology 2021: 210033.

7. Sudre CH, Murray B, Varsavsky T, et al. Attributes and predictors of long COVID. Nat Med 2021; 27: 626–31.

8. Ackermann M, Verleden SE, Kuehnel M, et al. Pulmonary Vascular Endothelialitis, Thrombosis, and Angiogenesis in Covid-19. New Engl J Med 2020; 383: 120–8.

9. Bösmüller H, Traxler S, Bitzer M, et al. The evolution of pulmonary pathology in fatal COVID-19 disease: an autopsy study with clinical correlation. Virchows Arch 2020; 477: 349–57.

10. Polak SB, Gool ICV, Cohen D, Thüsen JH von der, Paassen J van. A systematic review of pathological findings in COVID-19: a pathophysiological timeline and possible mechanisms of disease progression. Modern Pathol 2020; 33: 2128–38.

11. Pagnesi M, Baldetti L, Beneduce A, et al. Pulmonary hypertension and right ventricular involvement in hospitalised patients with COVID-19. Heart 2020; 106: 1324–31.

12. Si-Mohamed S, Chebib N, Sigovan M, et al. In vivo demonstration of pulmonary microvascular involvement in COVID-19 using dual-energy computed tomography. European Respir J 2020; 56: 2002608.

13. Ridge CA, Desai SR, Jeyin N, et al. Dual-Energy CT Pulmonary Angiography (DECTPA) Quantifies Vasculopathy in Severe COVID-19 Pneumonia. Radiology Cardiothorac Imaging 2020; 2: e200428.

14. Santamarina MG, Riscal DB, Beddings I, et al. COVID-19: What Iodine Maps From Perfusion CT can reveal—A Prospective Cohort Study. Crit Care 2020; 24: 619.

15. Grillet F, Behr J, Calame P, Aubry S, Delabrousse E. Acute Pulmonary Embolism Associated with COVID-19 Pneumonia Detected by Pulmonary CT Angiography. Radiology 2020; 296: 201544.

16. Jain A, Doyle DJ, Mangal R, et al. “Mosaic Perfusion Pattern” on Dual-Energy CT in COVID-19 Pneumonia: Pulmonary Vasoplegia or Vasoconstriction? Radiology Cardiothorac Imaging 2020; 2: e200433.

17. Evbuomwan O, Engelbrecht G, Bergman MV, Mokwena S, Ayeni OA. Lung perfusion findings on perfusion SPECT/CT imaging in non-hospitalized de-isolated patients diagnosed with mild COVID-19 infection. Egypt J Radiology Nucl Medicine 2021; 52: 144.

18. Rysz S, Al-Saadi J, Sjöström A, et al. COVID-19 pathophysiology may be driven by an imbalance in the renin-angiotensin-aldosterone system. Nat Commun 2021; 12: 2417.

19. Lucatelli P, Monte MD, Rubeis GD, et al. Did we turn a blind eye? The answer is simply there. Peripheral pulmonary vascular thrombosis in COVID-19 patients explains sudden worsening of clinical conditions. Imaging 2020; 12: 4–7.

20. Dhawan RT, Gopalan D, Howard L, et al. Beyond the clot: perfusion imaging of the pulmonary vasculature after COVID-19. Lancet Respir Medicine 2020; 9: 107–16.

21. Buonsenso D, Giuda DD, Sigfrid L, et al. Evidence of lung perfusion defects and ongoing inflammation in an adolescent with post-acute sequelae of SARS-CoV-2 infection. Lancet Child Adolesc Heal 2021; 5: 677–80.

22. Risse F, Eichinger M, Kauczor HU, Semmler W, Puderbach M. Improved visualization of delayed perfusion in lung MRI. Eur Radiol 2011; 77: 105–10.

23. Schiwek M, Triphan SMF, Biederer J, et al. Quantification of pulmonary perfusion abnormalities using DCE-MRI in COPD: comparison with quantitative CT and pulmonary function. Eur Radiol 2021: 1–12.

24. Raghu G, Remy-Jardin M, Myers JL, et al. Diagnosis of Idiopathic Pulmonary Fibrosis. An Official ATS/ERS/JRS/ALAT Clinical Practice Guideline. Am J Resp Crit Care 2018; 198: e44–68.

25. Kolb M, Collard HR. Staging of idiopathic pulmonary fibrosis: past, present and future. European Respir Rev 2014; 23: 220–4.

26. Bestall JC, Paul EA, Garrod R, Garnham R, Jones PW, Wedzicha JA. Usefulness of the Medical Research Council (MRC) dyspnoea scale as a measure of disability in patients with chronic obstructive pulmonary disease. Thorax 1999; 54: 581.

27. Daynes E, Gerlis C, Briggs-Price S, Jones P, Singh SJ. COPD assessment test for the evaluation of COVID-19 symptoms. Thorax 2020; 76: thoraxjnl-2020-215916.

28. Grimby G, Frändin K. On the use of a six-level scale for physical activity. Scand J Med Sci Spor 2018; 28: 819–25.

29. Anon. ATS Statement. Am J Resp Crit Care 2012; 166: 111–7.

30. Chetta A, Zanini A, Pisi G, et al. Reference values for the 6-min walk test in healthy subjects 20–50 years old. Resp Med 2006; 100: 1573–8.

31. Haitao T, Vermunt J, Abeykoon J, et al. COVID-19 and Sex Differences: Mechanisms and Biomarkers. Mayo Clin Proc 2020; 95: 2189–203.

32. Burnham KJ, Arai TJ, Dubowitz DJ, et al. Pulmonary perfusion heterogeneity is increased by sustained, heavy exercise in humans. J Appl Physiol 2009; 107: 1559–68.

33. Dehnert C, Risse F, Ley S, et al. Magnetic Resonance Imaging of Uneven Pulmonary Perfusion in Hypoxia in Humans. Am J Resp Crit Care 2006; 174: 1132–8.

34. Santamarina MG, Boisier D, Contreras R, Baque M, Volpacchio M, Beddings I. COVID-19: a hypothesis regarding the ventilation-perfusion mismatch. Crit Care 2020; 24: 395.

35. Heidenreich JF, Weng AM, Metz C, et al. Three-dimensional Ultrashort Echo Time MRI for Functional Lung Imaging in Cystic Fibrosis. Radiology 2020; 296: 191–9.

36. Simmons DH, Linde LM, Miller JH, O’Reilly RJ. Relation between lung volume and pulmonary vascular resistance. Circ 1961; 9: 465–71.

